# SARS-CoV-2 Heterogeneity by Ethnicity in Los Angeles

**DOI:** 10.1101/2021.05.10.21256955

**Authors:** Lao-Tzu Allan-Blitz, Fred Hertlein, Jeffrey D. Klausner

**Affiliations:** Brigham and Women’s Hospital, Boston, MA; Curative Inc. San Dimas, CA; Department of Preventative Medicine, Keck School of Medicine, University of Southern California

**Author notes:** **Corresponding Author Contact Information:** Lao-Tzu Allan-Blitz, Department of Medicine, Brigham and Women’s Hospital, 75 Francis Street, Boston, MA 02115, USA. **Alternative Author Information:** Dr. Jeffrey D. Klausner, Department of Preventative Medicine, Keck School of Medicine, University of Southern California, Los Angeles, CA 90033, USA. Compliance with Ethical Standards: Disclosures: Dr. Allan-Blitz has served as a consultant for Curative Inc. Dr. Klausner has served as the medical director of Curative Inc. during the observation period. Fred Hertlein has served as an employee of Curative Inc. during the observation period. Ethical Approval: This article does not contain any studies with human participants performed by any of the authors. The Mass General Brigham institutional review board deemed the analysis of de-identified data did not constitute human subjects’ research (2020P003530). No informed consent was necessary.

## Abstract

Recent studies have identified notable disparities in SARS-CoV-2 infection risk among ethnic minorities. We evaluated SARS-CoV-2 test results from individuals presenting for testing in Los Angeles between June-December, 2020. We calculated prevalence ratios for various employment categories. Among 518,914 test results, of which 295,295 (56.9%) were from individuals reporting Hispanic ethnicity, SARS-CoV-2 positivity was 16.5% among Hispanic individuals compared to 5.0% among non-Hispanic individuals (*p*-value<0.01). The prevalence ratios comparing Hispanic and non-Hispanic individuals was highest for members of the media (PR=6.7; 95% CI 4.3-10.4), government employees (PR=4.0; 95% CI 3.3-4.9), and agricultural workers (PR=4.0; 95% CI 3.2-5.0). Such heterogeneity warrants further investigation in order to develop targeted public health interventions towards specific drivers of SARS-CoV-2 transmission.

## Background

The impact of the SARS-CoV-2 pandemic has been disproportionately felt among Hispanic communities (1-3). The complex and interconnected structural and systemic factors that likely contribute to such disparities (4) are also likely driving the heterogeneity with which SARS-CoV-2 spreads (5, 6). Ironically, however, the broad business closures enforced in order to curb the pandemic may be less efficacious among minority and poorer communities, as such communities are less able to limit their mobility (e.g. working from home or limit within household contact due to dense housing and household occupancy) and are less likely to work in settings with regular testing or adequate ventilation, all of which may increase risk of SARS-CoV-2 infection (7). Even worse, such communities who have less financial security at the outset may feel the economic impacts of public health interventions most acutely (8, 9).

A new strategy, therefore, is desperately needed. In the wake of previous epidemics, targeted public health interventions have been made possible via a thorough understanding of factors that facilitate the transmission of disease. In response to the Human Immunodeficiency Virus (HIV) epidemic, for example, the HIV Prevention Trials Network has developed innumerable, novel, and cost-effective interventions that are tailored directly towards factors that drive infection (10). Similarly, targeted interventions may be possible for the SARS-CoV-2 pandemic if we are able to develop a more thorough understanding of the specific drives among sub-populations at highest risk.

We therefore aimed to evaluate the risk of SARS-CoV-2 infection associated with different employment categories among Hispanic compared to White individuals, in order to unwind the details of factors that contribute to the disparity in burden of infection. We performed that analysis among a community-based sample in Los Angles as Los Angeles county is among the most populous counties in the United States and one of the most diverse, with 48.6% of those surveyed in the national census identifying as Hispanic (11).

## Methods

We collected data from individuals presenting for SARS-CoV-2 polymerase chain reaction testing to one of 250 drive-through centers in Los Angeles county. Individuals presented between June-December, 2020. Specimens were collected via oral swabs and processed as has been previously reported (12). Data collected at the time of testing included age, gender, race and ethnicity, reported exposure to an individual known to be infected with SARS-CoV-2, report of symptoms (muscle aches, cough, fevers, diarrhea, nausea, headache, congestion, sore throat, anosmia, and shortness of breath), and type of employment from a pre-specified list. Laboratory data collected included human and viral cycle threshold values, date and time of testing.

We included data in our analysis for each individual test if there were no missing values for ethnicity, test result, and employment category. We then calculated the prevalence of SARS-CoV-2 test positivity by ethnicity and employment category as well as prevalence ratios and 95% confidence intervals (CI) for each employment category. The Mass General Brigham institutional review board deemed the analysis of de-identified data did not constitute human subjects’ research (2020P003530).

## Results

Overall, we analyzed 518,914 individual tests results, of which 296,052 (57.1%) were collected from women and 295,295 (56.9%) reported Hispanic ethnicity. The mean age of the population was 35 years (standard deviation 15.2). In all, 247,974 (47.8%) individuals reported any symptoms at the time of testing. SARS-CoV-2 positivity was 16.5% among Hispanic individuals compared to 5.0% among non-Hispanic individuals (*p*-value<0.01). The percent of individuals who reported symptoms at the time of testing among those who tested positive was comparable among Hispanic (70.9%) and non-Hispanic individuals (69.8%). The table shows SARS-CoV-2 positivity by employment category as well as prevalence ratios and 95% confidence intervals.

## Discussion

From a community-based sample of individuals presenting for testing in Los Angeles county, we identified a significant disparity in SARS-CoV-2 test positivity among Hispanic compared to non-Hispanic individuals. And though descriptive, we also identified notable heterogeneity in risk based on reported employment among Hispanic communities.

Many recent studies have demonstrated the disproportionate burden of SARS-CoV-2 infection suffered by minority races and ethnicities (1-4, 13-16). There are numerous factors that likely contribute to those findings, including the higher proportion of immigrant families without health insurance limiting the ability to seek care, language barriers impacting the quality of care received, financial insecurity resulting in need for potentially higher-risk employment or limiting the ability to work from home (17), and housing insecurity leading to denser household occupancy and thus increased risk for transmission. The tangled web of those factors as well as innumerable others creates significant challenges in developing public health strategies that optimally address the needs of such communities. In order to untangle such a web, we must first be able to precisely describe the specific scenarios that perpetuate those disparities. Testing centers can be used to gather detailed exposure data (e.g. exposure to gyms, places of worship, parks, museums, hospitals, nursing homes, overnight camps, hotels, beaches, movie theaters, bars, restaurants, and airports) as well as dates of those exposures in order to understand when and how the transmission chains occur. Furthermore, such data should be collected among individuals who test positive as well as negative, as the comparison of risk will be essential in evaluating which types of exposures truly drive transmission.

Our results demonstrated notably higher prevalence ratios for Hispanic individuals employed as members of the media, agriculture workers, and government personnel compared to non-Hispanics with similar employment. A recent study similarly found increased risk for SARS-CoV-2 positivity among kitchen workers of minority races (18). Beyond the known structural inequities that most likely contribute to such differences, there may be ongoing disparities in division of labor within the workforce of essential workers in specific roles that additionally compounds the heighted risk for transmission. For example, Hispanic members of the media may be more likely than non-Hispanic members to work in settings that place them at higher risk for SARS-CoV- 2 transmission and acquisition. One survey found that Hispanic individuals were more likely than White individuals to lose their job during the pandemic (19), which may suggest that the jobs held by Hispanic individuals were different in substance than their White counterparts, and thus reflect ongoing discrepant positions held by different ethnicities. We did not have sufficient data, however, to evaluate other confounding factors, thus further research is needed to more deeply evaluate such findings.

Our study was also limited in that all participants were tested in one geographic region, thus potentially limiting the generalizability of our results. However, given the diversity within Los Angeles (mirrored partially among our sample), and the fact that Los Angeles in particular is an important case study of the ongoing socioeconomic disparities surrounding the SARS-CoV-2 pandemic, we do not feel that limitation reduces the importance of our results. Our study is additionally limited in that we were unable to adjust our analysis based on socioeconomic status, and thus our results may be confounded, thus further research is warranted.

## Conclusion

Our results add further support to the existing body of research identifying notable disparities in SARS-CoV-2 test positivity among ethnic minorities. Furthermore, in an attempt to explore the heterogeneity in SARS-CoV-2 transmission within Hispanic compared to non-Hispanic communities we reported prevalence ratios for various employment categories, noting that several demonstrated significantly higher risk for infection. Further research is needed in order to precisely identify factors driving transmission within high-risk populations and thereby develop tailored public health interventions.

## Data Availability

The data are available upon request.

## Acknowledgements and Funding

The authors would like to acknowledge the city of Los Angeles. There was no funding for this study.

---

**Table:**
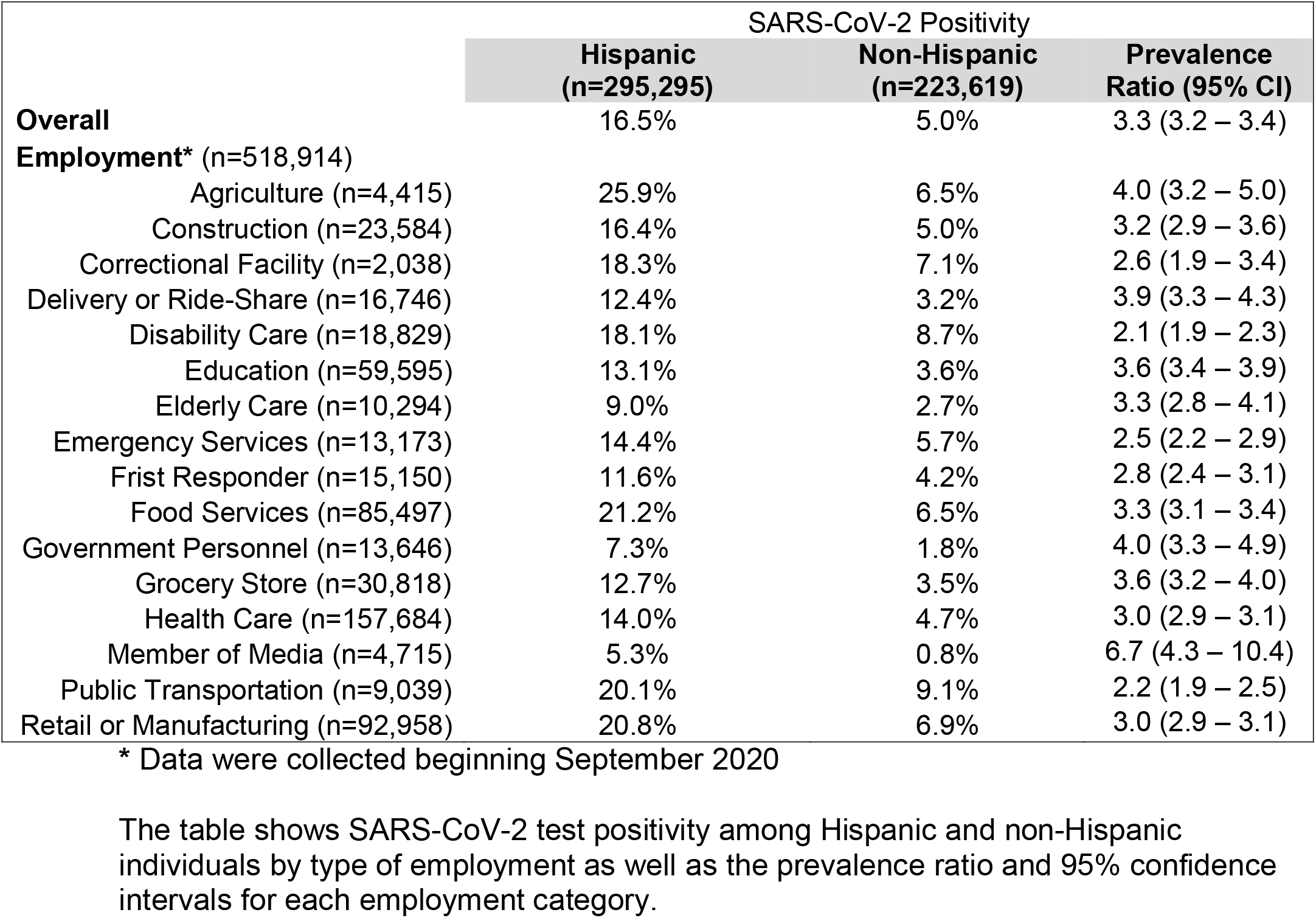
SARS-CoV-2 Positivity Among Hispanic and Non-Hispanic Individuals, Los Angeles June-December, 2020.

|  | SARS-CoV-2 Positivity |  |  |
| --- | --- | --- | --- |
|  | Hispanic<br>(n=295,295) | Non-Hispanic<br>(n=223,619) | Prevalence<br>Ratio (95% CI) |
| **Overall** | 16.5% | 5.0% | 3.3 (3.2 – 3.4) |
| **Employment\*** (n=518,914) |  |  |  |
| Agriculture (n=4,415) | 25.9% | 6.5% | 4.0 (3.2 – 5.0) |
| Construction (n=23,584) | 16.4% | 5.0% | 3.2 (2.9 – 3.6) |
| Correctional Facility (n=2,038) | 18.3% | 7.1% | 2.6 (1.9 – 3.4) |
| Delivery or Ride-Share (n=16,746) | 12.4% | 3.2% | 3.9 (3.3 – 4.3) |
| Disability Care (n=18,829) | 18.1% | 8.7% | 2.1 (1.9 – 2.3) |
| Education (n=59,595) | 13.1% | 3.6% | 3.6 (3.4 – 3.9) |
| Elderly Care (n=10,294) | 9.0% | 2.7% | 3.3 (2.8 – 4.1) |
| Emergency Services (n=13,173) | 14.4% | 5.7% | 2.5 (2.2 – 2.9) |
| Frist Responder (n=15,150) | 11.6% | 4.2% | 2.8 (2.4 – 3.1) |
| Food Services (n=85,497) | 21.2% | 6.5% | 3.3 (3.1 – 3.4) |
| Government Personnel (n=13,646) | 7.3% | 1.8% | 4.0 (3.3 – 4.9) |
| Grocery Store (n=30,818) | 12.7% | 3.5% | 3.6 (3.2 – 4.0) |
| Health Care (n=157,684) | 14.0% | 4.7% | 3.0 (2.9 – 3.1) |
| Member of Media (n=4,715) | 5.3% | 0.8% | 6.7 (4.3 – 10.4) |
| Public Transportation (n=9,039) | 20.1% | 9.1% | 2.2 (1.9 – 2.5) |
| Retail or Manufacturing (n=92,958) | 20.8% | 6.9% | 3.0 (2.9 – 3.1) |

\* Data were collected beginning September 2020
The table shows SARS-CoV-2 test positivity among Hispanic and non-Hispanic individuals by type of employment as well as the prevalence ratio and 95% confidence intervals for each employment category.

## Notes

### Funding Statement

There was no funding for this project.

### Author Declarations

This article does not contain any studies with human participants performed by any of the authors. The Mass General Brigham institutional review board deemed the analysis of de-identified data did not constitute human subjects research (2020P003530).

